# Reductions in stillbirths and preterm birth in COVID-19 vaccinated women: a multi-center cohort study of vaccination uptake and perinatal outcomes

**DOI:** 10.1101/2022.07.04.22277193

**Authors:** Lisa Hui, Melvin Barrientos Marzan, Daniel L. Rolnik, Stephanie Potenza, Natasha Pritchard, Joanne M. Said, Kirsten R Palmer, Clare L. Whitehead, Penelope M. Sheehan, Jolyon Ford, Ben W. Mol, Susan P. Walker

**Author notes:** **CORRESPONDING AUTHOR:** A/Prof Lisa Hui Dept of Perinatal Medicine, Level 3, Mercy Hospital for Women, 163 Studley Rd, Heidelberg, VIC 3084 Australia, fx. 61 3 8458 4504. **DISCLOSURES** LH has received research funding from Ferring Pharmaceuticals outside the scope of this work. BWM is a consultant for Guerbet and has received research grants from Guerbet and Merck. KRP has received consultancy fees from Janssen. DLR has received fees from Alexion for participation in advisory boards unrelated to this work. All other authors declare no competing interests. **PAPER PRESENTATION** The findings of this study were submitted as a free communication for the Annual Scientific Meeting of the Royal Australian and New Zealand College of Obstetricians and Gynaecologists at the Gold Coast, QLD, Australia in October 2022. **CONDENSATION** Stillbirths, spontaneous and iatrogenic preterm births significantly lower in COVID-19 vaccinated women.

## Abstract

**Background:** COVID-19 infection in pregnancy is associated with a higher risk of progression to severe disease, but vaccine uptake by pregnant women is hindered by persistent safety concerns. COVID-19 vaccination in pregnancy has been shown to reduce stillbirth, but its relationship with preterm birth is uncertain.

**Objective:** The aim of this study was to investigate the sociodemographic characteristics associated with vaccine uptake in Melbourne, Australia, and to compare perinatal outcomes by vaccination status.

**Study design:** Retrospective multicenter cohort study in Melbourne following the national recommendations for mRNA COVID-19 vaccination during pregnancy in June 2021. Routinely collected data from all 12 public maternity hospitals in Melbourne were extracted on births ≥ 20 weeks’ gestation from 1^st^ July 2021 to 31 March 2022. Maternal sociodemographic characteristics were analyzed from the total birth cohort. Perinatal outcomes were compared between vaccinated and unvaccinated women for whom weeks 20-43 of gestation fell entirely within the 9-month data collection period. The primary outcome was the rate of congenital anomaly in singleton infants ≥ 20 weeks’ gestation among women vaccinated during pregnancy. Secondary perinatal outcomes including stillbirth, preterm birth (spontaneous and iatrogenic), birthweight ≤ 3^rd^ centile, and newborn intensive care unit admissions were examined for singleton infants ≥ 24 weeks’ gestation without congenital anomalies. We calculated the adjusted odds ratio of congenital anomalies and perinatal outcomes among vaccinated versus unvaccinated women using inverse propensity score weighting regression adjustment with multiple covariates; p< 0.05 was considered statistically significant.

**Results:** Births from 32,536 women were analyzed: 17,365 (53.4%) were vaccinated and 15,171 (47.6%) were unvaccinated. Vaccinated women were significantly more likely to be older, nulliparous, non-smoking, not requiring an interpreter, of higher socioeconomic status, and vaccinated against pertussis and influenza. Vaccination status also varied by region of birth: compared with women born in Australia, women born in South and Eastern Europe, the Middle East, Africa and Oceania had lower adjusted odds of vaccination. There was no significant increase in the rate of congenital anomalies or birth weight ≤ 3^rd^ centile in vaccinated women. Vaccinated women were significantly less like to have an infant with a major congenital anomaly compared with the unvaccinated group (2.4% vs 3.0%, aOR 0.72, 95%CI 0.56-0.94, p=0.02). This finding remained significant even when the analysis was restricted to women vaccinated before 20 weeks’ gestation. Vaccinated women had a significantly lower rate of stillbirth (0.2% vs 0.8%, aOR 0.18, 95%CI 0.09-0.37, P < 0.001. Vaccination was associated with a significant reduction in total preterm births < 37 weeks (5.1% vs 9.2%, aOR 0.60, 95% CI 0.51-0.71, p< 0.001), spontaneous preterm birth (2.4% vs 4.0%, aOR 0.73 95% CI 0.56-0.96, p=0.02) and iatrogenic preterm birth (2.7% vs 5.2%, aOR 0.52, 95%CI 0.41-0.65, p< 0.001).

**Conclusions:** COVID-19 Vaccine coverage was significantly influenced by known social determinants of health, which is likely to influence the strong association between COVID-19 vaccination and lower risks of stillbirth and preterm birth. We did not observe any adverse impacts of vaccination on fetal growth or development.

**AT A GLANCE:** *Why was this study conducted?:*

⍰ COVID-19 infection in pregnancy is associated with a higher risk of progression to severe disease, but vaccine uptake by pregnant women is hindered by persistent safety concerns. COVID-19 vaccination in pregnancy has been shown to reduce stillbirth, but its relationship with preterm birth is uncertain.
⍰ Most of the published literature on COVID-19 vaccination in pregnancy have methodological limitations including fixed cohort bias and time-varying exposure.
⍰ We conducted this multicenter study to provide robust evidence on mRNA COVID-19 vaccination and perinatal outcomes including congenital anomalies, stillbirth, and preterm birth.

*What are the key findings?:*

⍰ The adjusted odds of stillbirth, preterm birth, and neonatal intensive care admission were significantly reduced among infants born to COVID-19 vaccinated women compared with unvaccinated women. COVID-19 vaccination during pregnancy was not associated with an increase in congenital anomalies.
⍰ Our results conclusively demonstrate a significant reduction in both spontaneous and iatrogenic preterm birth for vaccinated women
⍰ Vaccinated women were significantly more likely to be older, nulliparous, non-smoking, not requiring an interpreter, residing in a higher socioeconomic postcode, and vaccinated against pertussis and influenza. There were also significant differences in vaccination rates by region of birth.

*What does this study add to what is already known?:*

⍰ Our analysis confirmed a strong relationship between the COVID-19 mRNA vaccine and lower preterm births and stillbirths
⍰ In addition to its impact on reducing severe COVID-19 illness, vaccination may be a proxy for other biological and social determinants of health among our pregnant population.

## Introduction

Pregnancy is an independent risk factor for severe COVID-19 infection, associated with higher rates of maternal hospitalization, intensive care unit admission and mortality compared with nonpregnant adults.^1^ COVID-19 infection during pregnancy also increases the risk of serious adverse perinatal outcomes, including stillbirth, preterm birth, Caesarean birth, and preeclampsia.

An effective vaccine against SARS-CoV2 became available globally from early 2021,^2^ with the Australian roll-out commencing in March 2021.^3^ By the end of 2021, vaccine uptake in the eligible Australian population was 85 per 100.^4^ It is now well-established that the mRNA COVID-19 vaccine is safe and protective against severe COVID-19 disease in pregnancy,^5-8^ but this confidence was not present at the launch of the vaccine program due to the exclusion of pregnant women from the early clinical trials.^9^ It was not until 9^th^ June 2021 that Australian health authorities changed their advice on vaccination in pregnancy from cautiously recommending deferral to the postpartum period, to clearly recommending vaccination with an mRNA vaccine during pregnancy.^10^ The change in health advice was met with vaccine hesitancy among some pregnant Australian women, with common concerns including side effects of the vaccine for their newborn and a perception of inadequate safety information.^11^ Vaccination was available free of charge through a range of public vaccination centres, general practitioners, and within the antenatal clinics in some maternity services. However due to supply shortages, pregnant women could not readily access the COVID-19 vaccine until late July 2021.

A recent systematic review and meta-analysis of 23 studies confirmed the effectiveness of the COVID-19 vaccine in pregnancy, as well as showing a significant reduction in stillbirth.^12^ However, this study was inconclusive about the impact of COVID-19 vaccination on preterm birth. The meta-analysis of the six studies that reported on preterm birth showed a non-significant relative reduction in preterm birth of 10%. Only two studies accounted for time-dependent exposures, that is, included adjustment for the fact that the vaccination occurred at various gestations.^13^ Women vaccinated in late pregnancy are not at risk of very preterm birth, which skews the results towards fewer preterm births in the vaccinated group. Another limitation of the literature is failure to account for fixed cohort bias.^14^ Fixed cohort bias occurs in retrospective cohorts that are defined by date of birth using calendar start and end dates. This has the effect of introducing both left fixed truncation bias (start of data collection occurs at a fixed date, but at variable gestations for each included pregnancy) and right censoring bias (otherwise eligible subjects are not included because their birth occurs after the study end date). When the exposure rate (i.e. vaccination) is higher at the end of the study period, this will have the effect of overestimating the preterm birth rate in the exposed group as term vaccinated pregnancies that deliver after the end of the cohort date will not be captured.

In Melbourne, Australia, data collection on maternal COVID-19 vaccination status was mandated by the Department of Health for all births from 1^st^ July 2021. These vaccination data were collected by a collaboration of 12 Melbourne public maternity hospitals under a research protocol established to monitor the effect of the pandemic on clinical quality indicators.^15^ Data from the Collaborative Maternity and Newborn Dashboard for the COVID-19 pandemic (CoMaND) project were used here to (i) analyse the sociodemographic factors associated with COVID-19 vaccine uptake and (ii) assess the perinatal outcomes associated with vaccination in pregnancy including preterm birth, stillbirth and congenital anomalies.

## Materials and Methods

### Institutional Review Board approval

This study was given ethical approval from the Human Research Ethics Committees of Austin Health (Ref. HREC/64722/Austin-2020) and Mercy Health (Ref. 2020-031).

### Study population

We extracted routinely collected data on births ≥ 20 weeks from all 12 public maternity hospitals in Melbourne from 1^st^ July 2021 to 31^st^ March 2022. Approximately 80% of all hospital births in Melbourne occur in these study sites. Births in exclusively private hospitals and planned home births outside of publicly funded homebirth programs were not captured. However, women planning a private hospital or home birth would typically be transferred to a public hospital if they were at risk of preterm birth <31 weeks or required tertiary maternal-fetal medicine care.

### Outcome measures

Our analysis was performed with several denominator groups according to the outcomes of interest.

### 1) Total births cohort

#### Sociodemographic characteristics

We used the total number of women giving birth from 1^st^ July 2021 to 31 March 2022 to compare the characteristics of those with and without at least one dose of a COVID-19 vaccine before or during pregnancy. The characteristics included: maternal age, body mass index (BMI in kg/m^2^), smoking status, need for an English interpreter, maternal region of birth, socioeconomic status (assigned by residential postcode), parity, diabetes (none, gestational, pre-existing, not tested), influenza vaccination status, pertussis vaccination status, plurality, gestation at first antenatal visit ≤ 12 weeks, and geographical remoteness.

#### Vaccination status by the week of birth

To examine temporal patterns in the uptake of COVID-19 vaccine following the health authority recommendations, we measured the weekly proportion of births to women who had received one or more doses of the mRNA COVID-19 vaccine before or during pregnancy. Choropleth maps of socioeconomic index for areas (SEIFA) and vaccination rates were generated in Tableau™ (Version 2022.3).

### 2) Calculated LMP cohort

We used the calculated week of last menstrual period (cLMP), rather than week of birth, to define the vaccine-exposed and unexposed groups to ensure that any woman birthing from 20 to 43 weeks’ gestation would be captured in the birth data, thus avoiding ‘fixed cohort bias’. We subtracted the infant’s gestational age at birth in completed weeks from the week of birth to obtain the week of calculated LMP (cLMP). Using this cLMP, we defined a cohort containing women for whom weeks 20-43 of gestation would have occurred after mandatory vaccination status reporting commenced on 1st July 2021. This included women whose cLMP occurred during the 16 weeks from 11^th^ February 2021 to 31^st^ May 2021 inclusive (Figure 1).

**Figure 1.**
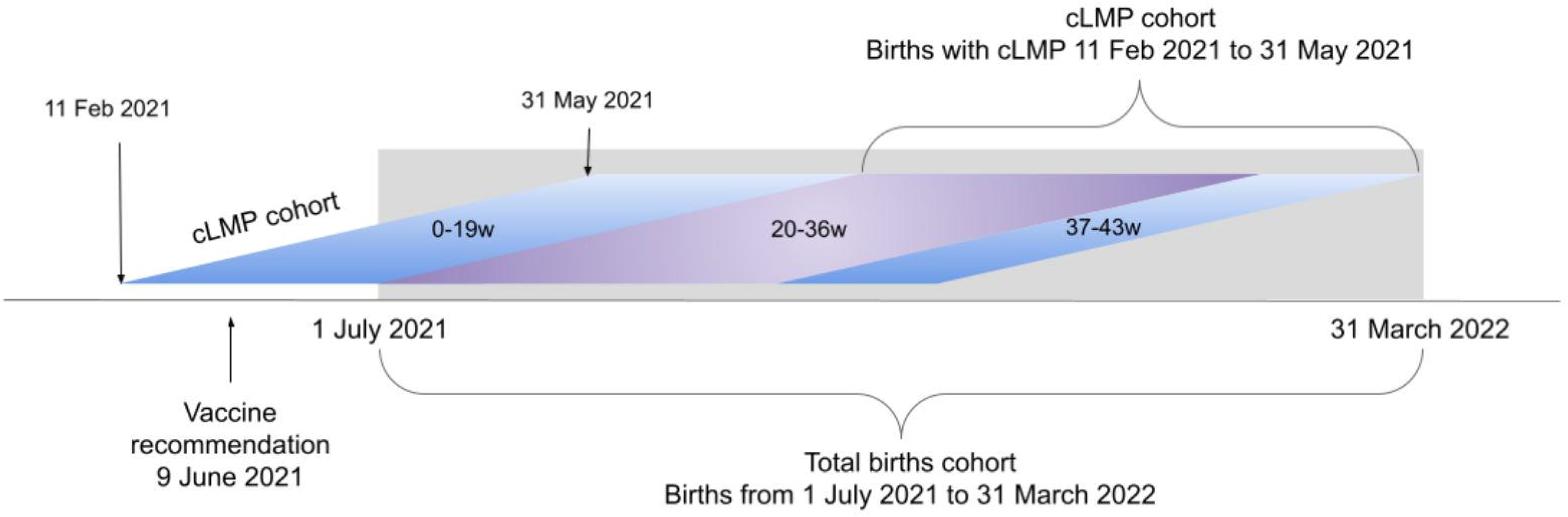
Total births and calculated last menstrual period (cLMP)cohorts

The pregnancies for which weeks 20-43 of gestation would have occurred during the birth data collection period (1^st^ July 2021 - 31 March 2022) were selected for the primary and secondary outcome analysis to avoid fixed cohort bias. Multifetal gestations and women who had preconception COVID-19 vaccination only were excluded from the singleton cLMP cohort.

#### Primary outcome: congenital anomalies ≥ 20 weeks

The primary outcome was the rate of major congenital anomalies in singleton infants ≥ 20 weeks’ gestation in women who had received at least one COVID-19 vaccination dose during pregnancy, including terminations of pregnancies.

#### Secondary outcomes: perinatal outcomes ≥ 24 weeks

Secondary outcomes were calculated from the singleton cLMP cohort, following exclusion of congenital anomalies, termination of pregnancies, and births < 24 weeks gestation.

Secondary outcomes were calculated using ‘all births’ denominator (live births and stillbirths)

1. Stillbirths: total, term, and preterm < 37 weeks
2. Preterm birth < 37 weeks: total, spontaneous, and iatrogenic. An iatrogenic birth was defined as any birth without spontaneous onset of labor (i.e. induced labor or no labor).
3. Fetal growth restriction (FGR): defined as birthweight ≤ 3^rd^ centile using Australian population sex-specific birth weight charts.^16^
4. Apgar score < 7 at five minutes
5. Special care nursery (SCN) admission
6. Neonatal intensive care unit (NICU) admission
7. Mode of birth: induction of labor, pre-labor Cesarean birth, Cesarean birth after labor onset, unassisted vaginal birth, instrumental vaginal birth (forceps/vacuum),
8. Born before arrival. This refers to the rate of planned hospital births that occur before arrival, including unplanned births at home, in transit, or other locations
9. Severe postpartum hemorrhage: estimated blood loss ≥ 1000ml
10. Iatrogenic birth for fetal compromise: total, ≥ 37 weeks, < 37 weeks. An iatrogenic birth was an induction of labor or Cesarean birth prior to labor onset. Indications for induction of labor and Cesarean births were coded according to the Australian Institute of Health and Welfare Metadata Online Registry, which defines fetal compromise as “suspected or actual fetal compromise, and intrauterine growth restriction”.^17^ Any documentation of suspected fetal growth restriction, antepartum abnormal cardiotocography, “fetal distress” (without labor), reduced fetal movements, oligohydramnios, abnormal umbilical artery Doppler studies, or placental insufficiency were included in this classification.

All secondary outcomes, other than the outcomes of congenital anomalies and stillbirths, were also calculated using live births as the denominator. Iatrogenic births for fetal compromise were only calculated using the live births denominator.

COVID-19 infections during pregnancy were collected and analyzed for the main outcomes of congenital anomalies, stillbirth and preterm birth according to vaccination status.

### Statistical analysis

No sample size calculation was performed as this was a cohort defined by the vaccination era period. Analyses of secondary outcomes were considered exploratory and no adjustments for multiple comparisons were made. Continuous variables in both determinants and safety analyses were presented with mean and standard deviation (SD). Categorical variables were presented as count and percentages. Statistical analyses were conducted using Stata 17 (StataCorp. 2021. Stata Statistical Software: Release 17. College Station, TX: StataCorp LLC), and two-sided p-values below 0.05 were considered statistically significant.

### Determinants of COVID-19 vaccination uptake

We performed multiple imputations by chained equation (MICE) to minimise the bias from missing data in our dataset and created five imputed datasets. Factors associated with antenatal COVID-19 vaccination were analyzed using multivariable Poisson regression with covariates selected based on subject matter knowledge. Results of the regression analyses were presented as Incident Rate Ratio (IRR). The hospital-specific rates of vaccination uptake were summarised and reported as forest plots.

### Perinatal outcomes

We used inverse-probability-weighted-regression-adjustment (IPTWRA) of Stata ‘teffects’ suite of commands to balance the baseline difference in the population by COVID-19 vaccination status. The IPTWRA also accounts for the missing data. We adjusted for the following covariates: maternal age, body mass index (BMI) at first antenatal visit, maternal region of birth, need for interpreter (proxy indicator for primary language and categorised as yes or no), parity, socioeconomic status (using quintiles of SocioEconomic Index for Areas, SEIFA for maternal postcode), diabetes status (non, gestational, pre-existing, not tested), and smoking in pregnancy status. These covariates were chosen using subject matter knowledge. Adjusted analyses were performed using logistic regression and presented as Odds Ratio s(OR).

#### Sensitivity analyses

We performed unadjusted and adjusted logistic regression analysis without IPTWRA to check the consistency of the directions of associations between COVID-19 vaccination and maternal and perinatal outcomes.

We performed a sensitivity analysis for congenital anomalies by excluding women who were first vaccinated ≥ 20 weeks, as the biologically relevant period of exposure for teratogenesis is in early pregnancy.

To examine the impact of time-varying exposure and the ‘healthy vaccinee” effect on preterm birth and stillbirth, we performed a Cox regression analysis excluding women who received their first dose of COVID-19 vaccine ≥ 24 weeks gestation and adjusted for pertussis vaccination status. We used Kaplan-Meier curves to plot the cumulative hazard of the outcomes of interest. The proportionality of the hazards of control and exposed cohorts was tested using Schoenfeld residuals.

## Results

There were 33,018 infants born to 32,536 women during the study period. The numbers of inclusions and exclusions for the various cohorts are shown in the study flowchart in Figure 2. The weekly percentage of births to vaccinated women are shown in Figure 3. By the end of the study period 85% of women giving birth had received at least one dose of the COVID-19 vaccine before or during pregnancy (Figure 3).

**Figure 2.**
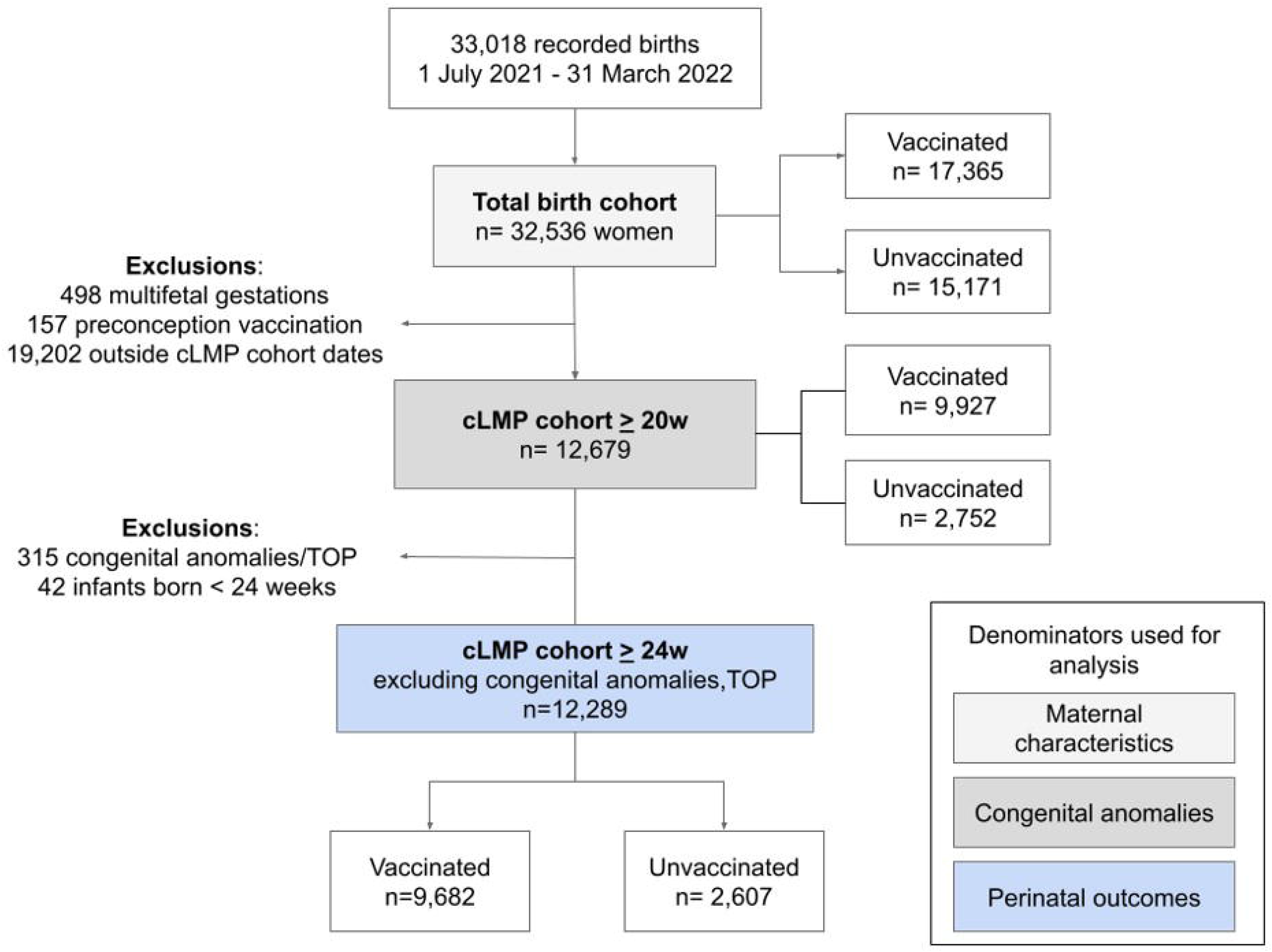
Flowchart of study cohorts

**Figure 3.**
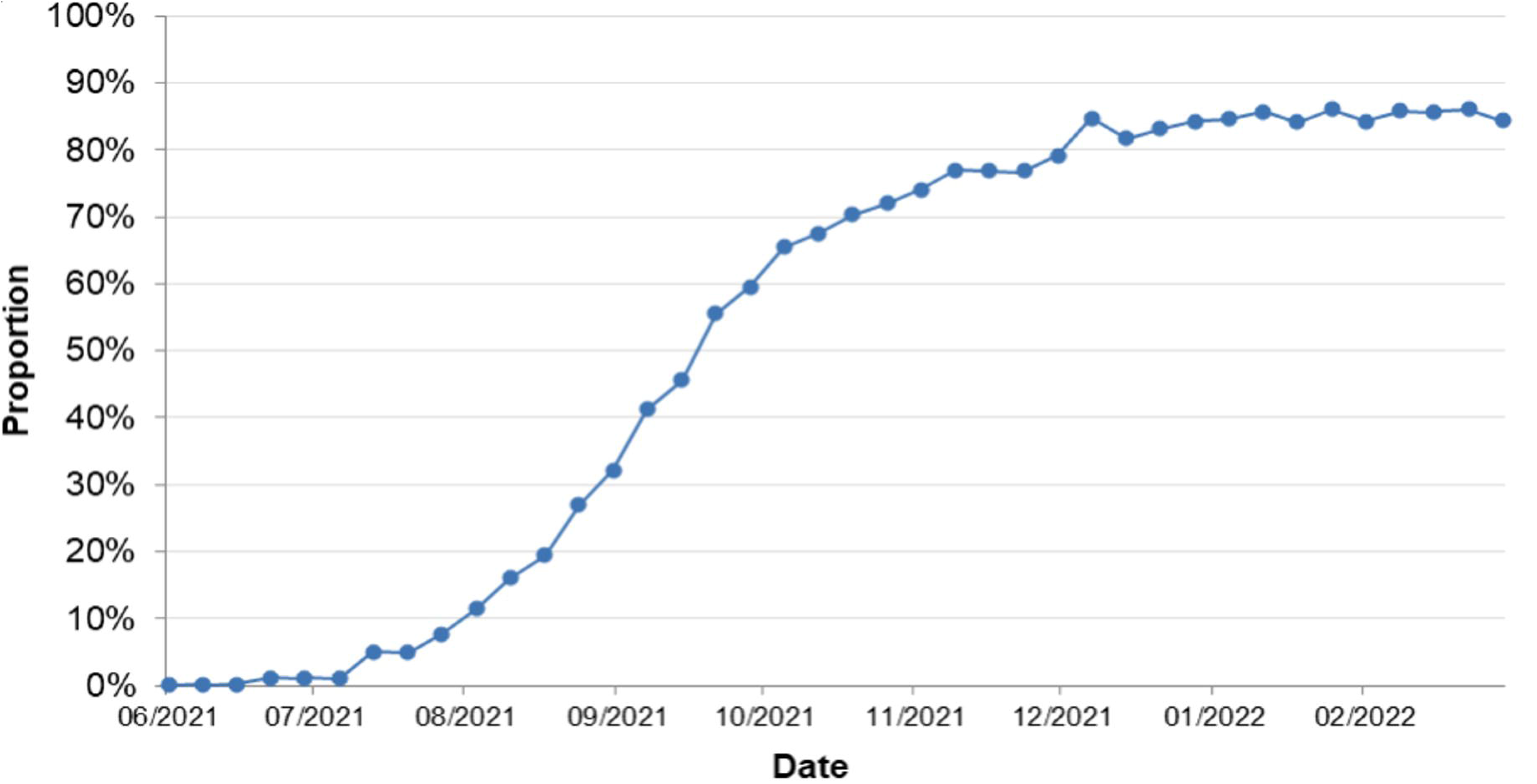
Weekly percentage of births to vaccinated women July 2021 to March 2022

The weekly percentage of births to women with COVID-19 infection during pregnancy are shown in Figure 4, representing a cumulative total of 1078 infected women. By the end of March 2022, 13% of women giving birth had experienced a COVID-19 infection during their pregnancy.

**Figure 4.**
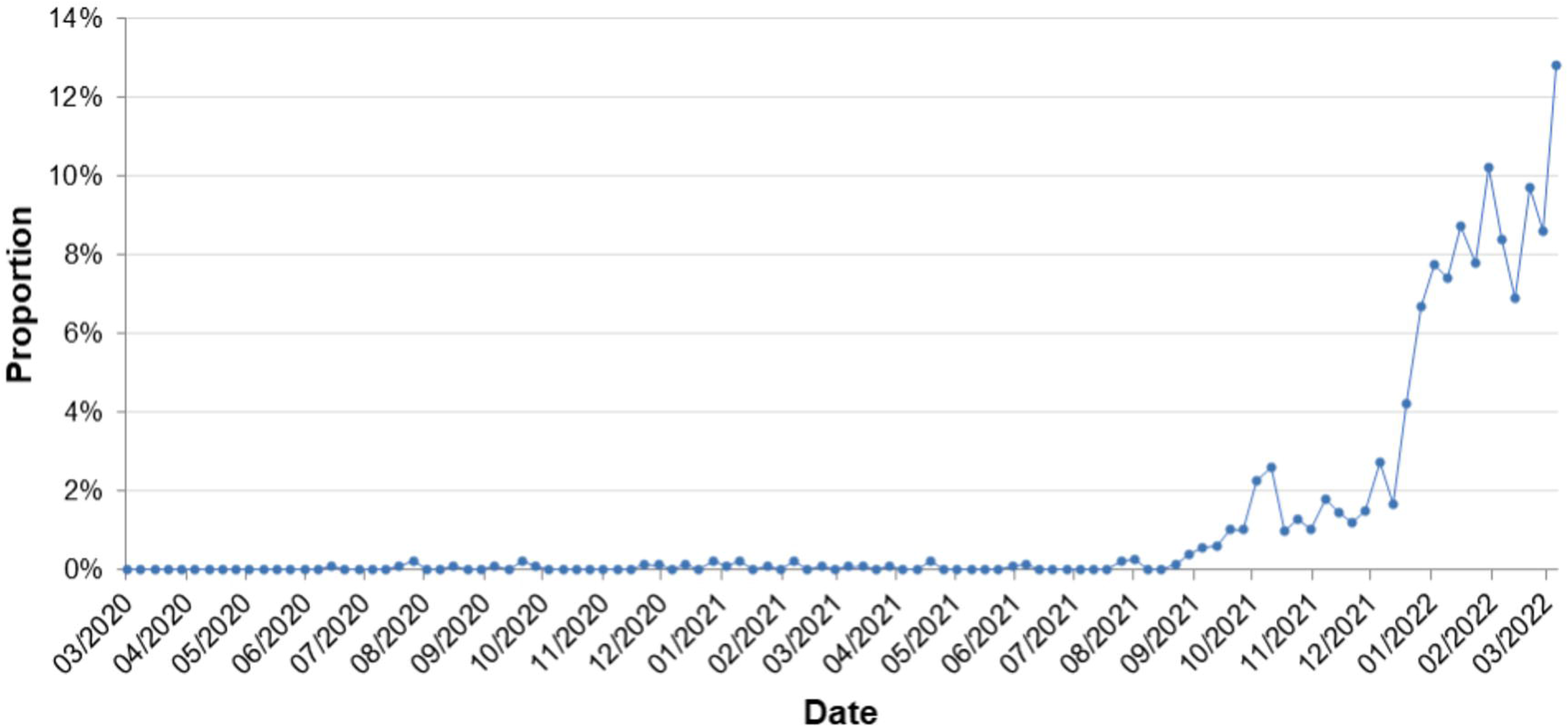
Weekly rate of births to women with COVID-19 infection during pregnancy

### Sociodemographic characteristics of vaccinated vs unvaccinated groups

Of the 32,536 women giving birth during the 9-month study period, 17,365 (53.4%) had received at least one dose of the mRNA COVID-19 vaccine and 15,171 (46.6%) had not. The characteristics of vaccinated and unvaccinated groups are shown in Table 1. Vaccinated women were significantly older, and more often nulliparous, non-smoking, not requiring an interpreter, of higher socioeconomic status, and vaccinated against pertussis and influenza. Vaccination status also varied by region of birth. Women from the Middle East, North Africa, Southern and Eastern Europe, and Oceania were less likely to be vaccinated than Australian-born women; women born in Southeast Asia were significantly more likely to be vaccinated. Choropleth maps of postcodes showing socioeconomic status and vaccination coverage are shown in Figure 5. The Forest plot of vaccination coverage by hospitals are provided in Figure 6.

**Table 1.** Maternal characteristics by COVID-19 vaccination status among total births cohort

| Maternal characteristics | Vaccinated<br>N=17,365 |  | Unvaccinated<br>N=15,171 |  | Multiple imputed dataset |  |  |
| --- | --- | --- | --- | --- | --- | --- | --- |
|  | n | % | n | % | Denominator | IRR* | P-value |
| Maternal age group, years |  |  |  |  |  |  |  |
| <25 | 1,477 | 8.5 | 1,915 | 12.6 | 32,536 | 0.91 | 0.005 |
| 25-29 | 3,223 | 18.6 | 3,377 | 22.3 |  | Ref |  |
| 30-34 | 7,224 | 41.6 | 5,823 | 38.4 |  | 1.12 | <0.001 |
| 35-39 | 4,487 | 25.8 | 3,327 | 21.9 |  | 1.18 | <0.001 |
| ≥ 40 | 954 | 5.5 | 729 | 4.8 |  | 1.19 | <0.001 |
| Parity |  |  |  |  |  |  |  |
| 0 | 7,783 | 44.8 | 6,259 | 41.3 | 32,535 | 1.09 | <0.001 |
| 1 or more | 9,582 | 55.2 | 8,911 | 58.7 |  | Ref |  |
| Plurality |  |  |  |  |  |  |  |
| Singleton | 17,102 | 98.5 | 14,936 | 98.5 | 32,536 | Ref |  |
| Multifetal pregnancy | 263 | 1.5 | 235 | 1.6 |  | 1.00 | 0.96 |
| BMI Categories |  |  |  |  |  |  |  |
| <18 | 193 | 1.2 | 173 | 1.2 | 30,824 | 0.99 | 0.90 |
| 18-24 | 7,651 | 46.3 | 6,477 | 45.3 |  | Ref |  |
| 25-29 | 4,892 | 29.6 | 4,322 | 30.0 |  | 1.00 | 0.97 |
| 30-34 | 2,228 | 13.5 | 1,950 | 13.6 |  | 1.03 | 0.27 |
| 35-39 | 947 | 5.7 | 860 | 6.0 |  | 1.04 | 0.26 |
| >=40 | 603 | 3.7 | 528 | 3.7 |  | 1.08 | 0.092 |
| Smoking in pregnancy |  |  |  |  |  |  |  |
| Non-smokers | 16,883 | 97.2 | 14,270 | 94.1 | 32,536 |  |  |
| Smokers | 482 | 2.8 | 901 | 5.9 |  | 0.71 | <0.001 |
| Region of Birth |  |  |  |  |  |  |  |
| Americas | 317 | 1.8 | 209 | 1.4 | 32,352 | 1.05 | 0.37 |
| Australia | 8,916 | 51.7 | 7,898 | 52.3 |  | Ref |  |
| North Africa and Middle East | 661 | 3.8 | 829 | 5.5 |  | 0.89 | 0.005 |
| Northeast Asia | 600 | 3.5 | 447 | 3.0 |  | 1.01 | 0.76 |
| Northwest Europe | 616 | 3.6 | 373 | 2.5 |  | 1.09 | 0.050 |
| Oceania | 527 | 527.0 | 633 | 4.2 |  | 0.90 | 0.018 |
| Southeast Asia | 1,525 | 8.8 | 1,042 | 6.9 |  | 1.12 | <0.001 |
| Southern and Central Asia | 3,426 | 19.9 | 2,758 | 18.3 |  | 1.03 | 0.17 |
| Southern and Eastern Europe | 249 | 1.4 | 390 | 2.6 |  | 0.71 | <0.001 |
| Sub-Saharan Africa | 426 | 2.5 | 510 | 3.4 |  | 0.89 | 0.022 |
| Interpreter required |  |  |  |  |  |  |  |
| Non-required | 16,645 | 95.9 | 14,413 | 95.0 | 32,536 |  |  |
| Required | 720 | 4.2 | 758 | 5.0 |  | 0.96 | 0.27 |
| SEIFA quintile |  |  |  |  |  |  |  |
| 1 - Most disadvantaged | 3,288 | 19.1 | 3,551 | 23.7 | 32,181 | 0.94 | 0.005 |
| 2 | 2,466 | 14.4 | 2,223 | 14.8 |  | 1.00 | 0.96 |
| 3 | 4,283 | 24.9 | 3,899 | 26.0 |  | Ref |  |
| 4 | 3,980 | 23.2 | 3,124 | 20.8 |  | 1.05 | 0.039 |
| 5 - Most advantaged | 3,159 | 18.4 | 2,208 | 14.7 |  | 1.07 | 0.007 |

| Remoteness of residence |  |  |  |  |  |  |  |
| --- | --- | --- | --- | --- | --- | --- | --- |
| Regional | 304 | 1.8 | 358 | 2.4 | 32,536 | Ref |  |
| Metropolitan Location | 17,061 | 98.3 | 14,813 | 97.6 |  | 0.93 | 0.19 |
| Gestation at first antenatal visit |  |  |  |  |  |  |  |
| ≤ 12 weeks | 12,252 | 70.6 | 10,656 | 70.2 | 32,536 | Ref |  |
| >12 weeks | 5,113 | 29.4 | 4,515 | 29.8 |  | 0.99 | 0.58 |
| Antenatal Influenza Vaccine |  |  |  |  |  |  |  |
| Non-vaccinated | 3,815 | 22.2 | 4,899 | 32.9 | 32,081 | Ref |  |
| Vaccinated | 13,362 | 77.8 | 10,005 | 67.1 |  | 1.24 | <0.001 |
| Antenatal Pertussis Vaccine |  |  |  |  |  |  |  |
| Non-vaccinated | 1,415 | 8.2 | 3,114 | 20.8 | 32,280 | Ref |  |
| Vaccinated | 15,860 | 91.8 | 11,891 | 79.3 |  | 1.74 | <0.001 |
| Diabetes Mellitus |  |  |  |  |  |  |  |
| None | 13,012 | 75.0 | 11,555 | 76.2 | 32,527 | Ref |  |
| Gestational diabetes mellitus | 4,088 | 23.6 | 3,299 | 21.8 |  | 1.03 | 0.10 |
| Pre-existing diabetes mellitus | 197 | 1.1 | 143 | 0.9 |  | 1.10 | 0.17 |
| Not tested for gestational diabetes mellitus | 64 | 0.4 | 169 | 1.1 |  | 0.61 | <0.001 |

BMI, body mass index; SD, standard deviation; SEIFA, socioeconomic index for areas; w, weeks;

**Figure 5.**
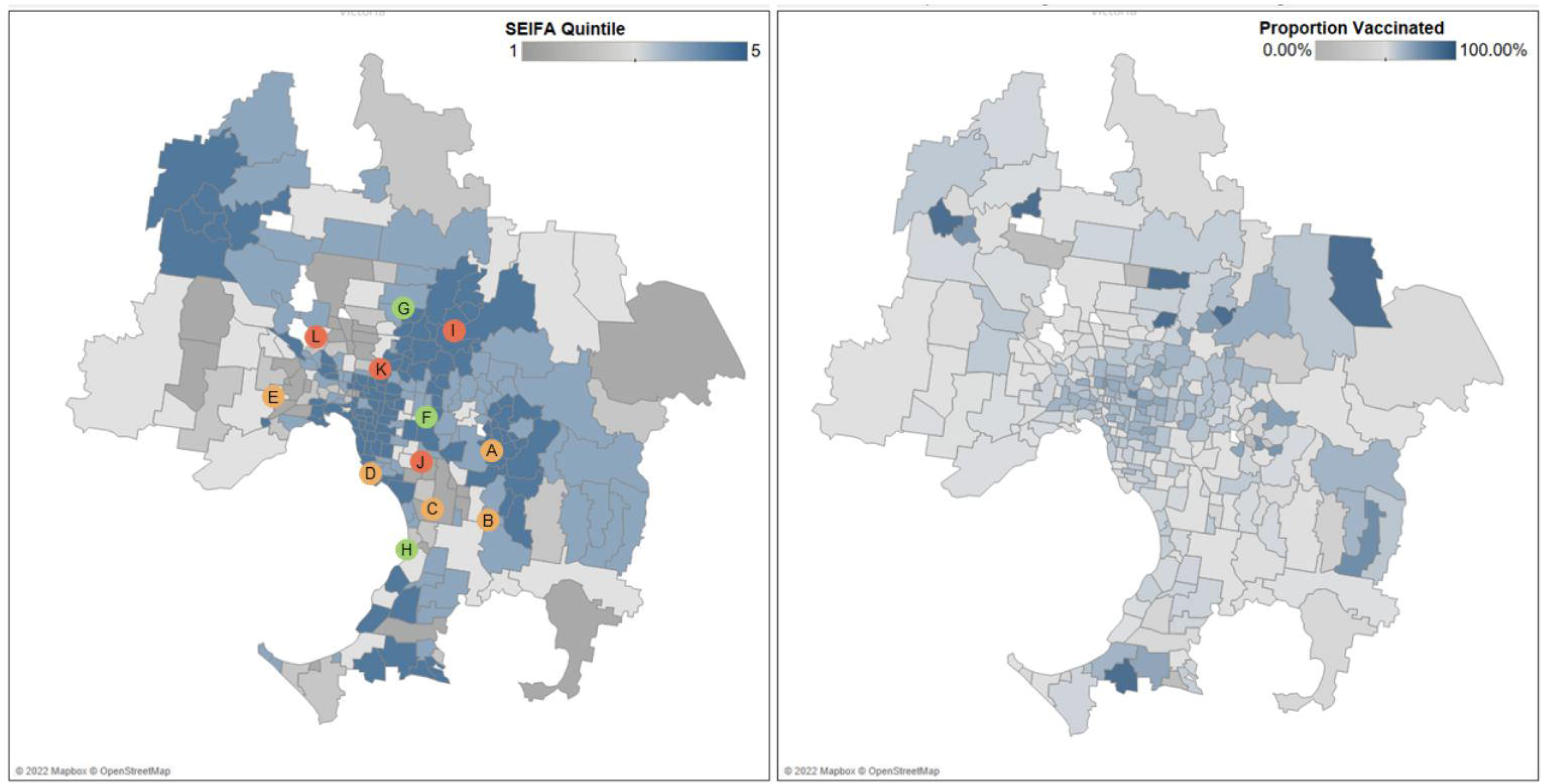
Socioeconomic index for areas and maternal vaccination coverage by postcode The participating hospitals are indicated by the lettered circles with their corresponding vaccination rates displayed in Figure 6.

**Figure 6.**
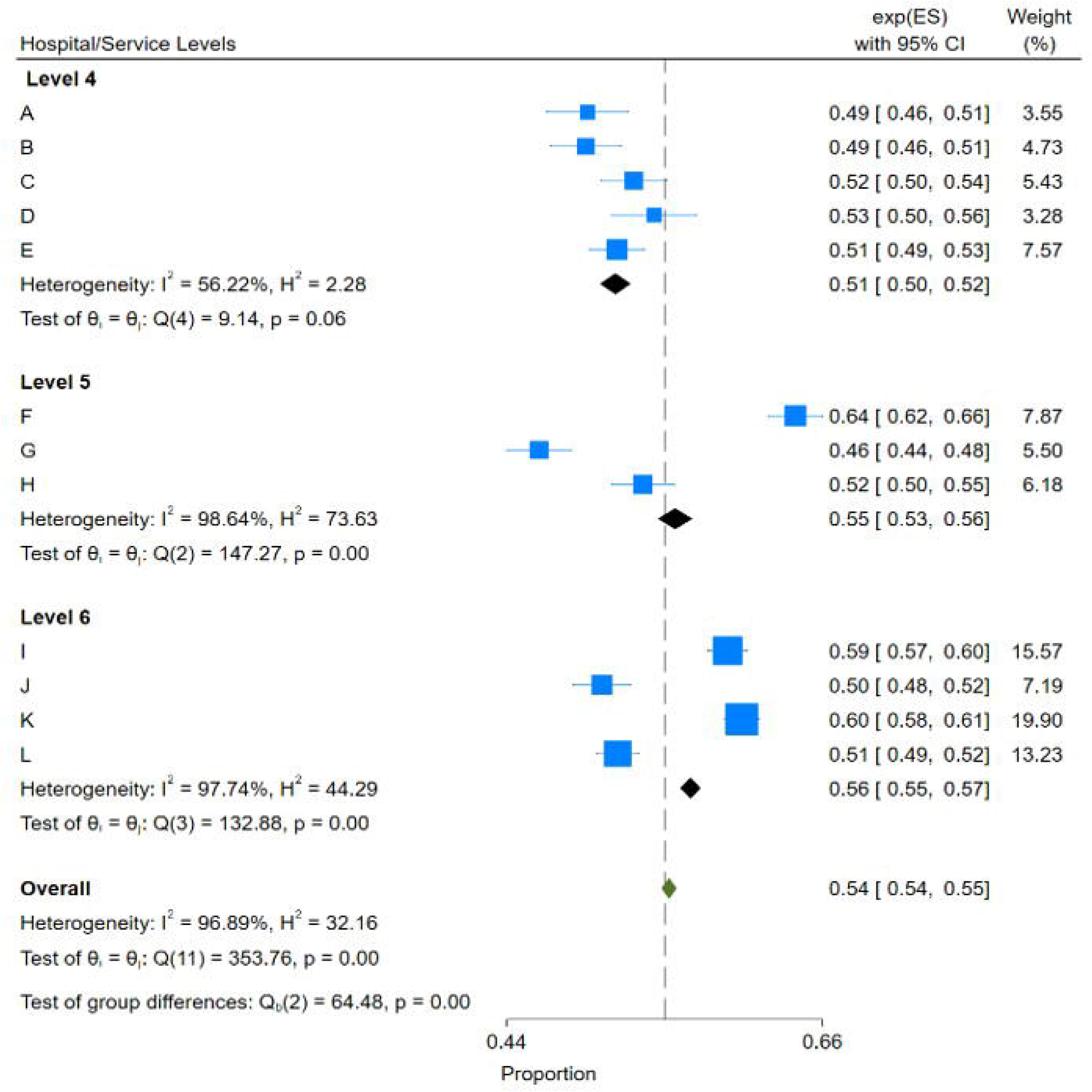
Forest plots of vaccination coverage by hospital* *The hospital level data is grouped here by maternity service level. Level 6 maternity services provide regional/statewide specialised care for high risk pregnancies, including extremely preterm births, as well as local care for all women and babies; Level 5 services care for normal to moderate risk pregnancies, and manage labor and birth from 31 weeks gestation; Level 4 services that provide local care for women and babies at normal and moderate risk, including planned births from 34 weeks gestation. The geographical location of each hospital are indicated in Figure 5.

### Perinatal outcomes in the cLMP cohort

After exclusions, 12,679 births remained in the cLMP cohort (Figure 2). Of these, 9927 had received at least one dose of the COVID-19 vaccination during pregnancy. A documented gestation at first and second vaccine dose was available for 98.3% and 92.7% of the vaccinated groups respectively. The median gestational age at the first dose was 24 weeks (interquartile range 19-28 weeks) and 28 w (interquartile range 24-32 weeks) for the second dose (Figure 7).

**Figure 7.**
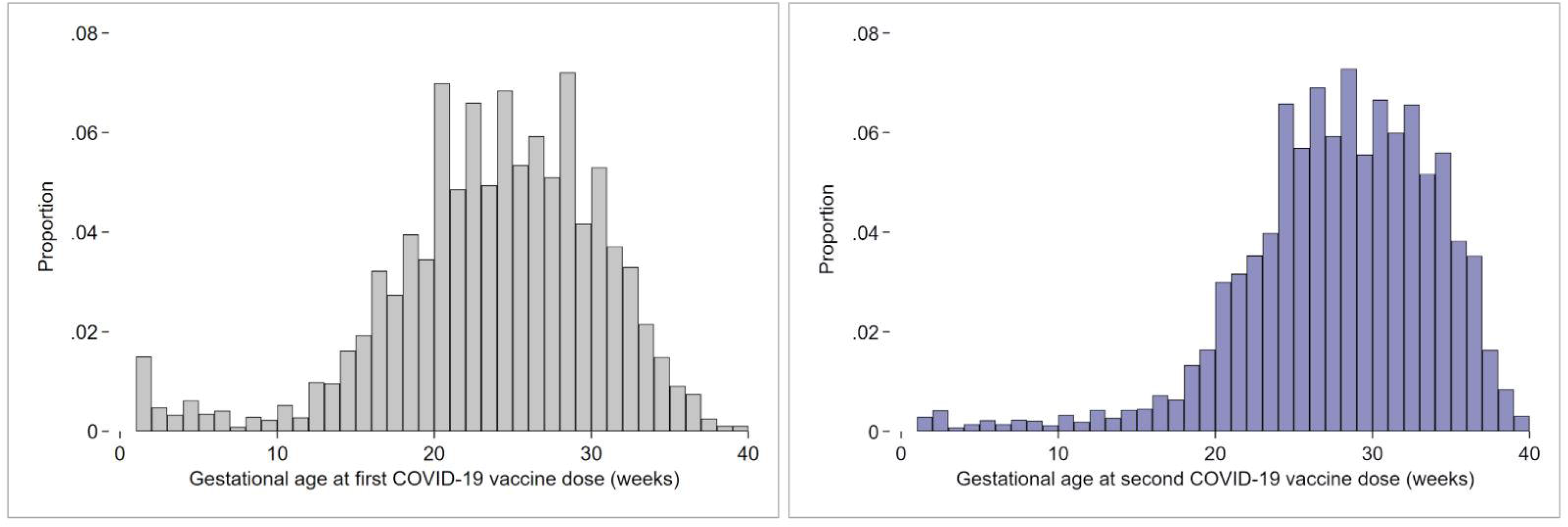
Timing of first and second COVID-19 vaccine doses during pregnancy* *Calculated on the singleton cLMP cohort > 20 weeks gestation

The primary and secondary outcomes with ‘all births’, and ‘live births’ denominators are presented in Tables 2 and 3 respectively.

**Table 2.** Primary and secondary outcomes among all births (live births and stillbirths)

| Primary and secondary outcomes | Vaccinated |  | Unvaccinated |  | Unadjusted Odds Ratio (IPTWRA) |  |  |  | Adjusted Odds Ratio (IPTWRA) |  |  |  |
| --- | --- | --- | --- | --- | --- | --- | --- | --- | --- | --- | --- | --- |
|  | n | (%) | n | (%) | OR | L | U | P value | aOR | L | U | P value |
| Singleton cLMP cohort ≥ 20 weeks | N=9927 | | N=2752 | | | | | | | | | |
| Congenital anomalies | 236 | 2.4 | 83 | 3.0 | 0.79 | 0.62 | 1.01 | 0.06 | 0.72 | 0.56 | 0.94 | 0.02 |
| Singleton cLMP cohort excluding congenital anomalies, TOP, and births < 24 weeks | N=9682 |  | N=2607 |  |  |  |  |  |  |  |  |  |
| Stillbirths |  |  |  |  |  |  |  |  |  |  |  |  |
| Stillbirths | 15 | 0.2 | 21 | 0.8 | 0.19 | 0.10 | 0.37 | 0.00 | 0.18 | 0.09 | 0.37 | <0.001 |
| Stillbirths >37 weeks | 6 | 0.1 | 3 | 0.1 | 0.54 | 0.13 | 2.15 | 0.38 | 0.38 | 0.09 | 1.55 | 0.18 |
| Stillbirths <37 weeks | 9 | 0.1 | 18 | 0.7 | 0.13 | 0.06 | 0.30 | 0.00 | 0.12 | 0.05 | 0.31 | <0.001 |
| Preterm birth <37 weeks |  |  |  |  |  |  |  |  |  |  |  |  |
| Total | 495 | 5.1 | 239 | 9.2 | 0.56 | 0.48 | 0.65 | 0.00 | 0.60 | 0.51 | 0.71 | <0.001 |
| Spontaneous | 234 | 2.4 | 104 | 4.0 | 0.61 | 0.48 | 0.76 | 0.00 | 0.73 | 0.56 | 0.96 | 0.02 |
| Iatrogenic | 261 | 2.7 | 135 | 5.2 | 0.52 | 0.42 | 0.64 | 0.00 | 0.52 | 0.41 | 0.65 | <0.001 |
| Newborn Outcomes |  |  |  |  |  |  |  |  |  |  |  |  |
| Fetal growth restriction | 237 | 2.5 | 63 | 2.4 | 1.01 | 0.77 | 1.33 | 0.93 | 1.07 | 0.79 | 1.44 | 0.68 |
| SCN admission | 907 | 9.4 | 281 | 10.8 | 0.87 | 0.77 | 0.99 | 0.03 | 0.92 | 0.80 | 1.06 | 0.27 |
| NICU admission | 236 | 2.4 | 106 | 4.1 | 0.60 | 0.48 | 0.75 | 0.00 | 0.70 | 0.53 | 0.91 | 0.01 |
| Apgars < 7 at 5 minutes | 170 | 1.8 | 80 | 3.1 | 0.57 | 0.44 | 0.74 | 0.00 | 0.72 | 0.51 | 1.01 | 0.06 |
| Intrapartum outcomes |  |  |  |  |  |  |  |  |  |  |  |  |
| Induction of labor | 3589 | 37.1 | 835 | 32 | 1.16 | 1.09 | 1.23 | 0.00 | 1.11 | 1.04 | 1.18 | 0.001 |
| Unassisted Vaginal Births | 4,738 | 48.9 | 1,378 | 52.9 | 0.93 | 0.89 | 0.97 | 0.00 | 0.98 | 0.94 | 1.02 | 0.38 |
| Instrumental Vaginal Births | 1,533 | 15.8 | 338 | 13.0 | 1.22 | 1.09 | 1.36 | 0.00 | 1.08 | 0.96 | 1.21 | 0.22 |
| Cesarean birth - no onset of labor | 1,829 | 18.9 | 529 | 20.3 | 0.93 | 0.85 | 1.02 | 0.11 | 0.90 | 0.81 | 0.99 | 0.04 |
| Cesarean birth - after onset of labor | 1,583 | 16.4 | 360 | 13.8 | 1.18 | 1.06 | 1.32 | 0.00 | 1.07 | 1.07 | 1.21 | 0.27 |
| Born before arrival | 62 | 0.6 | 27 | 1.0 | 0.62 | 0.39 | 0.97 | 0.04 | 0.81 | 0.50 | 1.31 | 0.39 |
| Severe PPH ≥ 1000ml | 806 | 8.3 | 205 | 7.9 | 1.06 | 0.91 | 1.23 | 0.45 | 0.97 | 0.83 | 1.13 | 0.72 |

\*adjusted for maternal age, metropolitan vs regional residence, smoking status, need for interpreter, body mass index, region of birth, socioeconomic index for postcodes, diabetes mellitus, parity, infant sex, gestation at first antenatal visit
aOR, adjusted odds ratio; cLMP, calculated last menstrual period; IPTWRA, inverse proportionality treatment weighted regression adjustment; L, lower limit of 95% confidence interval; SCN, special care nursery; NICU, neonatal intensive care unit; OR, odds ratio; PPH, postpartum hemorrhage; TOP, termination of pregnancy; U, Upper limit of 95% confidence interval

**Table 3.** Secondary outcomes among live births

| Singleton cLMP cohort excluding congenital anomalies, TOP, births < 24 weeks and stillbirths | Vaccinated<br>N=9667 |  | Unvaccinated<br>N=2586 |  | Unadjusted Odds Ratio (IPTWRA) |  |  |  | Adjusted Odds Ratio* (IPTWRA) |  |  |  |
| --- | --- | --- | --- | --- | --- | --- | --- | --- | --- | --- | --- | --- |
|  | n | (%) | n | (%) | OR | L | U | P value | aOR | L | U | P value |
| Preterm birth <37 weeks |  |  |  |  |  |  |  |  |  |  |  |  |
| Total | 486 | 5.0 | 221 | 8.6 | 0.59 | 0.50 | 0.69 | <0.001 | 0.64 | 0.54 | 0.76 | <0.001 |
| Spontaneous | 230 | 2.4 | 102 | 3.9 | 0.60 | 0.48 | 0.76 | <0.001 | 0.72 | 0.55 | 0.94 | 0.017 |
| Iatrogenic | 256 | 2.7 | 119 | 4.6 | 0.58 | 0.47 | 0.71 | <0.001 | 0.58 | 0.46 | 0.74 | <0.001 |
| Newborn Outcomes |  |  |  |  |  |  |  |  |  |  |  |  |
| Fetal growth restriction | 236 | 2.4 | 62 | 2.4 | 1.02 | 0.77 | 1.34 | 0.899 | 1.08 | 0.79 | 1.46 | 0.63 |
| SCN admission | 907 | 9.4 | 281 | 10.9 | 0.86 | 0.76 | 0.98 | 0.023 | 1.08 | 0.80 | 1.06 | 0.27 |
| NICU admission | 236 | 2.4 | 106 | 4.1 | 0.60 | 0.48 | 0.75 | <0.001 | 0.69 | 0.53 | 0.91 | 0.01 |
| Apgars < 7 at 5 minutes | 155 | 1.6 | 60 | 2.3 | 0.69 | 0.51 | 0.93 | 0.013 | 1.39 | 0.51 | 1.01 | 0.06 |
| Intrapartum outcomes |  |  |  |  |  |  |  |  |  |  |  |  |
| Induction of labor | 3,584 | 37.1 | 820 | 31.7 | 1.17 | 1.10 | 1.24 | <0.001 | 1.12 | 1.05 | 1.19 | 0.001 |
| Cesarean birth - no onset of labor | 1,826 | 18.9 | 526 | 20.3 | 0.93 | 0.85 | 1.01 | 0.094 | 0.90 | 0.81 | 0.99 | 0.04 |
| Unassisted Vaginal Births | 4,728 | 48.9 | 1,360 | 52.6 | 0.93 | 0.89 | 0.97 | 0.001 | 0.99 | 1.06 | 1.03 | 0.50 |
| Instrumental Vaginal Births | 1,531 | 15.8 | 338 | 13.1 | 1.21 | 1.09 | 1.35 | 0.001 | 1.07 | 0.95 | 1.20 | 0.27 |
| Cesarean birth - after onset of labor | 1,583 | 16.4 | 360 | 13.9 | 1.18 | 1.06 | 1.31 | 0.003 | 0.30 | 0.94 | 1.20 | 0.30 |
| Born before arrival | 61 | 0.6 | 26 | 1.0 | 0.63 | 0.40 | 0.99 | 0.046 | 0.83 | 0.51 | 1.35 | 0.45 |
| Severe PPH ≥ 1000ml | 805 | 8.3 | 205 | 7.9 | 1.05 | 0.91 | 1.22 | 0.512 | 0.96 | 0.83 | 1.13 | 0.64 |
| Iatrogenic births for fetal compromise |  |  |  |  |  |  |  |  |  |  |  |  |
| Iatrogenic birth for fetal compromise | 1,853 | 19.17 | 451 | 17.44 | 0.46 | 0.34 | 0.64 | 0.00 | 0.49 | 0.34 | 0.70 | <0.001 |
| Iatrogenic birth for fetal compromise ≥ 37 weeks | 1,751 | 18.11 | 392 | 15.16 | 1.19 | 1.08 | 1.32 | 0.00 | 1.11 | 1.00 | 1.24 | 0.040 |
| Iatrogenic birth for fetal compromise < 37 weeks | 102 | 1.06 | 59 | 2.28 | 0.46 | 0.34 | 0.64 | 0.00 | 0.49 | 0.34 | 0.70 | <0.001 |

\*adjusted for maternal age, metropolitan vs regional residence, smoking status, need for interpreter, body mass index, region of birth, socioeconomic index for postcodes, diabetes mellitus, parity, infant sex, gestation at first antenatal visit aOR, adjusted odds ratio; cLMP, calculated last menstrual period; IPTWRA, inverse proportionality treatment weighted regression adjustment; L, lower limit of 95% confidence interval; SCN, special care nursery; NICU, neonatal intensive care unit; OR, odds ratio; PPH, postpartum hemorrhage; TOP, termination of pregnancy; U, Upper limit of 95% confidence interval

### Primary outcome

#### Congenital anomalies

The rate of major congenital anomalies was significantly lower in the vaccinated group than in the unvaccinated group (2.4% vs 3.0%, aOR 0.72, 95%CI 0.56-0.94, p=0.02) (Table 2). This did not remain statistically significant in the sensitivity analysis that excluded women vaccinated ≥ 20 weeks’ gestation, though the direction of the trend was similar (2.6% vs 3.0%, aOR 0.80, 95%CI 0.57-1.13, p=0.21) (Table 4).

**Table 4.** Sensitivity analysis for congenital anomalies, stillbirths, preterm births, and nursery admissions among all births

| Outcomes | Vaccinated |  | Unvaccinated |  | Unadjusted Odds Ratio (IPTWRA) |  |  |  | Adjusted Odds Ratio*(IPTWRA) |  |  |  |
| --- | --- | --- | --- | --- | --- | --- | --- | --- | --- | --- | --- | --- |
|  |  |  |  |  | OR | L | U | P value | aOR | L | U | P value |
| Singleton cLMP cohort vaccinated < 20 weeks | N=2442 |  | N=2668 |  |  |  |  |  |  |  |  |  |
|  | n | % | n | % |  |  |  |  |  |  |  |  |
| Congenital anomalies | 66 | 2.6 | 83 | 3.0 | 0.87 | 0.63 | 1.20 | 0.400 | 0.80 | 0.57 | 1.13 | 0.21 |
| Singleton cLMP cohort vaccinated < 24 weeks excluding congenital anomalies, terminations of pregnancy and births < 24 weeks | N=2442 |  | n=4665 |  |  |  |  |  |  |  |  |  |
|  | n | % | n | % |  |  |  |  |  |  |  |  |
| Stillbirths |  |  |  |  |  |  |  |  |  |  |  |  |
| Stillbirths | 6 | 0.1 | 21 | 0.8 | 0.16 | 0.06 | 0.39 | <0.001 | 0.17 | 0.07 | 0.44 | <0.001 |
| Stillbirths >37 weeks | 2 | 0.0 | 3 | 0.1 | 0.37 | 0.06 | 2.23 | 0.28 | 0.22 | 0.04 | 1.39 | 0.11 |
| Stillbirths <37 weeks | 4 | 0.1 | 18 | 0.7 | 0.12 | 0.04 | 0.37 | <0.001 | 0.15 | 0.05 | 0.47 | 0.001 |
| Preterm birth < 37 weeks |  |  |  |  |  |  |  |  |  |  |  |  |
| Total | 231 | 5.0 | 239 | 9.2 | 0.54 | 0.45 | 0.64 | <0.001 | 0.58 | 0.48 | 0.71 | <0.001 |
| Spontaneous | 114 | 2.4 | 104 | 4.0 | 0.61 | 0.47 | 0.80 | <0.001 | 0.73 | 0.73 | 0.99 | 0.046 |
| Iatrogenic | 117 | 2.5 | 135 | 5.2 | 0.73 | 0.54 | 0.99 | 0.046 | 0.48 | 0.36 | 0.63 | <0.001 |
| Newborn outcomes |  |  |  |  |  |  |  |  |  |  |  |  |
| SCN admission | 422 | 9.1 | 281 | 10.9 | 0.84 | 0.73 | 0.97 | 0.016 | 0.90 | 0.77 | 1.05 | 0.19 |
| NICU admission | 134 | 2.9 | 106 | 4.1 | 0.71 | 0.55 | 0.91 | 0.006 | 0.81 | 0.61 | 1.09 | 0.17 |

\*adjusted for maternal age, metropolitan vs regional residence, smoking status, need for interpreter, body mass index, region of birth, socioeconomic index for postcodes, diabetes mellitus, parity, infant sex, gestation at first antenatal visit
aOR, adjusted odds ratio; cLMP, calculated last menstrual period; IPTWRA, inverse proportionality treatment weighted regression adjustment; L, lower limit of 95% confidence interval; SCN, special care nursery; NICU, neonatal intensive care unit; OR, odds ratio; PPH, postpartum hemorrhage; TOP, termination of pregnancy; U, Upper limit of 95% confidence interval

### Secondary outcomes

The vaccinated group had a significantly lower rate of stillbirth than the unvaccinated group (0.2% vs 0.8%, aOR 0.18, 95%CI 0.09-0.37, P < 0.001)(Table 2). When stratified by gestational age, this difference was statistically significant only for preterm stillbirths (0.1% vs 0.7%, aOR 0.12, 95%CI 0.05-0.31, p < 0.001). This finding remained robust in the sensitivity analysis (Table 4).

The vaccinated group also had a significantly lower rate of preterm birth (5.1% vs 9.2%, aOR 0.60, 95%CI 0.51-0.71, p< 0.001), which was significant for both spontaneous preterm birth (2.4% vs 4.0%, aOR 0.73, 95% 0.56-0.96, P=0.02) and iatrogenic preterm birth (2.7% vs 5.2%, aOR 0.52, 95%CI 0.41-0.65, P< 0.001). These findings remained robust in the sensitivity analysis (Table 4). The hazard ratio plots for preterm birth and stillbirths that excluded women vaccinated ≥ 24 weeks and adjusted for pertussis vaccination status are shown in Figure 8.

**Figure 8.**
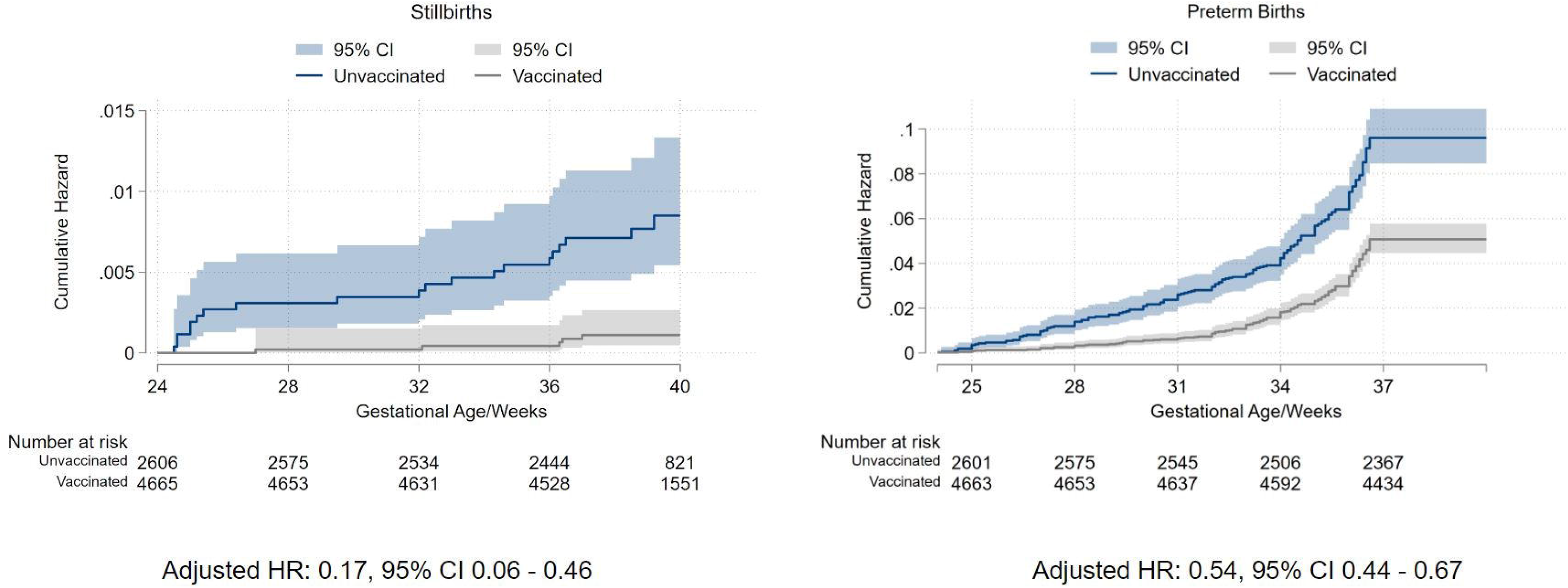
Hazard ratio plots for stillbirth and preterm birth for women vaccinated before 24 weeks gestation* *Plots were generated using the multiple imputed dataset and included adjustment for pertussis vaccination status

There was also a significant reduction in admissions to NICU in the vaccinated group (2.4% vs 4.1%, aOR 0.70, 95%CI 0.53-0.91, P= 0.007). This trend to lower NICU admissions was observed in the sensitivity analysis, but with a p value > 0.05.

There was no difference in the rate of severe fetal growth restriction between the vaccinated and unvaccinated groups. The vaccinated group had a significantly higher rate of induction of labor and iatrogenic births for fetal compromise at term gestation, but significantly lower rate of iatrogenic birth for fetal compromise at preterm gestations and lower rate of pre-labor Cesarean births (Table 3). There was no significant difference in other birth outcomes, such as PPH, vaginal births, or births before arrival to hospital.

#### Subgroup analysis of stillbirth and preterm birth by COVID-19 infection during pregnancy and vaccination status

There were 1,078 women with a COVID-19 infection during pregnancy in the total study cohort; of these, 518 women were in the cLMP cohort (Table 5). The crude OR of COVID-19 infection was significantly lower in the vaccinated group (3.6% vs 5.9%, OR 0.59, 95%CI 0.49-0.71, p < 0.0001). There was also a significant reduction in preterm birth among COVID-19 infected women who had been vaccinated during pregnancy (2.3% vs 7.0%, OR 0.32, 95% CI 0.12-0.80, p=0.015) (Table 5). The number of infected cases was too low to allow adjustment for covariates.

**Table 5.** Selected outcomes among women with documented COVID-19 infection by vaccination status

| Outcomes | Vaccinated |  | Unvaccinated |  | Unadjusted Odds |  |  |  |
| --- | --- | --- | --- | --- | --- | --- | --- | --- |
|  | N=9927 |  | N=2752 |  | OR | L | U | P value |
|  | count | (%) | count | (%) |  |  |  |  |
| Singleton cLMP cohort with COVID-19 infection | N=355 |  | N=163 |  |  |  |  |  |
| Congenital anomalies | 9 | 2.5 | 4 | 2.5 | 1.0 | 0.3 | 3.4 | 0.96 |
| Singleton cLMP cohort excluding congenital anomalies, TOP, and births < 24 weeks with COVID-19 infection | N=346 |  | N=158 |  |  |  |  |  |
| Stillbirths | 1 | 0.3 | 2 | 1.3 | 0.2 | 0.0 | 2.5 | 0.23 |
| Total preterm birth < 37 weeks | 8 | 2.3 | 11 | 7.0 | 0.3 | 0.1 | 0.8 | 0.02 |
| Spontaneous preterm birth | 1 | 0.3 | 4 | 2.5 | 0.1 | 0.0 | 1.0 | 0.05 |
| Iatrogenic preterm birth | 7 | 2.0 | 7 | 4.4 | 0.5 | 0.2 | 1.3 | 0.14 |

cLMP, calculated last menstrual period; L, lower limit of 95% confidence interval; OR, odds ratio; TOP, termination of pregnancy; U, Upper limit of 95% confidence interval

### Comment

#### Principal findings

Our multicenter cohort study confirms the safety and benefits of COVID-19 vaccination in pregnancy and adds new insights into the relationship between vaccination and preterm birth. Vaccinated women did not have higher rates of congenital anomalies or fetal growth restriction, but had significantly lower rates of stillbirth and preterm birth compared with unvaccinated women. Of note, our study shows that COVID-19 vaccination during pregnancy is associated with significantly lower rates of both spontaneous and iatrogenic preterm birth, even after excluding women vaccinated ≥ 24 weeks’ gestation.

#### Results in the Context of What is Known

Our finding on congenital anomalies provides additional reassuring safety data on COVID-19 vaccination in pregnancy. We have strengthened the existing evidence base by showing that there was no increase in the rate of congenital anomalies for women vaccinated before 20 weeks’ gestation, when the risk of teratogenesis would be expected to be higher. The significant reduction in stillbirths is in keeping with previously published studies, but our results are remarkable for the magnitude of the effect size. Our aOR of 0.18 is much lower than any of the seven individual studies included in the recent meta-analysis, which reported ORs of 0.50-1.50.^12^ This effect was even more profound in the preterm stillbirths, where vaccination was associated with a 1 in 1,000 of risk preterm stillbirth compared with 7 in 1,000 without vaccination.

That vaccination reduces stillbirth makes biological sense given the strong association between severe COVID-19 infection and an increased risk of stillbirth.^1^ However, as an observational study, we can only infer a causal relationship between vaccination and fewer stillbirths, presumably mediated by the significant reduction in the odds of COVID-19 infection. However, the burden of COVID-19 infection in the unvaccinated cohort did not appear high enough to account for the entire effect of vaccination on stillbirths. Although we controlled for multiple covariates, and performed a sensitivity analysis on pertussis vaccination status, we speculate that other relevant unmeasured factors associated with COVID-19 vaccination persist, such as health literacy or healthcare-seeking behavior (the ‘healthy vaccinee’ bias).^18^ The significantly higher rates of term induction of labor and better adherence to universal gestational diabetes screening in the vaccinated group suggest important variations in obstetric care that may have contributed to the difference in perinatal outcomes.

Our study provides the strongest evidence to date to support the association between COVID-19 vaccination and reduction in iatrogenic and spontaneous preterm births. The reduction in iatrogenic preterm birth may be due to prevention of complications directly related to COVID-19 infection such as preeclampsia, or severe maternal respiratory morbidity, while the reduction in spontaneous preterm birth may be related to avoiding the systemic inflammation associated with COVID-19 infection or other non-specific immune effects. Furthermore, unmeasured confounders may also be responsible for the reduction in preterm births, as discussed above for stillbirths.

The sociodemographic characteristics of our unvaccinated cohort mirror the findings from the UK showing variation in vaccination uptake by socioeconomic status, age, and region of birth.^19^ In multicultural Melbourne, public health messaging to non-English speaking communities had notable flaws, including incorrect and out-of-date translations of vaccine communications.^21^ This may have contributed to a lack of access to accurate vaccine safety information and a lack of trust in the health authorities among certain ethnic groups.

#### Clinical implications

Our data gives us strong evidence to confidently endorse the safety of the mRNA vaccine for our local pregnant population and to promote the benefits of reduced stillbirth and preterm birth. Our vaccine coverage of 85% at the end of March 2022 is an achievement that surpasses those recorded in the United States and the United Kingdom (70% and 60% respectively in 2022).^19,21^ However, the substantial regional variation in vaccine coverage within Melbourne (Figure 6) reflects the socioeconomic and cultural diversity of our population. A sensitive, multifaceted approach in partnership with community leaders is required to improve public health communication and remove barriers to vaccination uptake.^22^

#### Research implications

The significant reduction in stillbirth and preterm birth raises research questions that may lead to improved care outside the pandemic. Biological and social factors responsible for the reduction in spontaneous preterm birth should be investigated, including nonspecific inflammatory-mediated mechanisms for preterm birth and the mitigating effects of vaccination for COVID-19 and other diseases.

It is also likely that vaccination status is a proxy for unmeasured social determinants of health, in addition to the well-established demographic factors included in our analysis. This ‘healthy vaccinee bias’ may reflect the quality of relationship between a pregnant woman and her health care provider, as personal recommendation by a health care practitioner remains one of the most important influences on vaccine uptake.^11^ It is also correlated with trust in the government.^23^ Understanding these confounders and developing new metrics to capture them would aid future studies of vaccination in pregnancy and perinatal outcomes.

#### Strengths and limitations

Our large multicenter cohort of all public hospitals captured 80% of total births in Melbourne, including all hospitals that were designated by the government to care for COVID-19 pregnant inpatients. The major strengths of our cohort are its size, timeliness of data collection, avoidance of common methodological biases, and detailed individual patient level data including gestation at first vaccine dose.

#### Limitations

This was a retrospective study using routinely collected maternity data. Only data on first and second doses of COVID-19 vaccine were collected by hospitals during the study period, so the uptake of the third dose by pregnant women is unknown. The 1078 women with COVID-19 infection in the total cohort should be considered a minimum estimate of our total COVID-19 caseload. We do not have individual medical record data to determine whether the stillbirths and preterm births in the unvaccinated cohort were the direct result of acute COVID-19 disease. We await the outcomes from the CHOPAN registry for detailed local information on pregnancy outcomes for women with COVID-19.^24^

Our maternity data collection only includes birth outcomes from 20 weeks’ gestation. We therefore cannot comment on risk of miscarriage after COVID-19 vaccination, although other large studies have already provided reassuring data on this.^25,26^ We acknowledge that our findings on the risk of congenital anomalies are limited by the lack of data on early pregnancy. The lower rate of congenital anomalies observed in the vaccinated group may be a result of differences in utilization of aneuploidy screening, ultrasound scans, and termination of pregnancy for fetal abnormality before 20 weeks. There may have also been a true reduction in the prevalence of congenital anomalies in the vaccinated group due to differences in periconceptional health behaviors, such as the use of folic acid supplementation, pre-conception optimization of medical conditions, avoidance of teratogens, and dietary quality.^27^

## Conclusions

COVID-19 vaccination during pregnancy is strongly associated with lower risks of stillbirth and preterm birth, without any adverse impacts on fetal growth or development. Vaccine coverage was not distributed equally in our population and was significantly influenced by known social determinants of health. Our findings highlight both the achievements of our vaccination program and the challenges for maternity care in ensuring equity of health outcomes across our population.

## Data Availability

Non-identifiable individual participant data is available on request to the Austin Health Human Research Ethics Committee and the Mercy Health Human Research Ethics Committee. The study investigators may contribute aggregate and non-identifiable individual patient data to national and international collaborations whose proposed use of the data has been ethically reviewed and approved by an independent committee and following signing of an appropriate research collaboration agreement with the University of Melbourne.

## Contributors

LH, SW, JS, CW, DR, BM, KP and SP conceived of the original collaboration, LH designed, coordinated, and acquired funding for the study; LH and MM drafted the data analysis plan, and all authors edited and/or approved the analysis plan; LH, SP, JS, DR, CW, PS, JF, MM collected the primary data; MM performed the data cleaning and coding; LH, MM and DR performed the data analysis; MM and LH created the tables and figures; LH and MM wrote the primary manuscript; all authors reviewed and edited the draft manuscript for scientific content and approved the final submitted manuscript.

## Acknowledgements

The health services and individual hospitals contributing to the Collaborative Maternity and Newborn Dashboard for COVID-19 pandemic are:

⍰ Mercy Health (Mercy Hospital for Women, Werribee Mercy Hospital)
⍰ The Royal Women’s Hospital, The Women’s at Sandringham
⍰ Monash Health (Monash Medical Center, Casey Hospital, Dandenong Hospital)
⍰ Northern Health (The Northern Hospital)
⍰ Western Health (Joan Kirner Women’s and Children’s Hospital)
⍰ Eastern Health (Box Hill Hospital, The Angliss Hospital)
⍰ Peninsula Health (Frankston Hospital)

We thank the health service data managers and research midwives (Tania Fletcher, Lynn Rigg, Michelle Knight, Eleanor Johnson, Abby Monaghan, Pauline Hamilton, Roshanee Perera) for their assistance with primary data collection and Mr Andrew Goldsachs for assistance with coding of the indications for iatrogenic births.

